# Anatomical, Functional, and Hemodynamic Determinants of Increased Pulmonary Capillary Wedge Pressure V-Wave Amplitude

**DOI:** 10.1101/2023.11.16.23298656

**Authors:** Mikhail Torosoff, Ciril Khorolsky, Riju Banerjee, Neil Yager

## Abstract

**Introduction:** Increased pulmonary capillary wedge pressure (PCWP) V-wave amplitude has been traditionally associated with significant mitral regurgitation (MR). However, increased PCWP V-waves may also occur in patients with elevated left ventricular end-diastolic pressure (LVEDP) or with dilated left atrium (LA). Interactions between MR, increased LVEDP, and LA dilatation in the genesis of increased PCWP V-waves were not well investigated.

**Material and Methods:** It was a single-center cohort study of 153 consecutive hemodynamically stable patients referred for invasive hemodynamic evaluation of dyspnea. Patients with significant valvular disease other than MR, acute coronary syndromes (ACS), hemodynamic instability, pericardial disease, and mechanical ventilation or mechanical LV support were excluded.

**Results:** Study cohort included 41% males, 68.4+/-11.9 years old, 60% were on beta-adrenergic antagonists and 69.3% were on afterload reduction therapy. PCWP V-wave amplitude was significantly increased in patients with moderate or more severe MR (p<0.001), dilated LA (p=0.002), with a trend towards increased V-wave amplitude in elevated >12 mmHg LVEDP (p=0.117). There were no significant differences in age, gender, prevalence of LV dysfunction, other co-morbidities, and/or E/e’ ratio according to the MR degree, LA volume, and LVEDP. In patients categorized according to moderate or more severe MR, dilated LA, and increased >12 mmHg LVEDP - the presence of any two of these abnormal parameters was associated with 50-65% prevalence of elevated V-waves and reached 100% when all three were parameters were abnormal (p<0.002).

**Conclusions:** In euvolemic patients undergoing elective invasive hemodynamic evaluation of dyspnea, elevated PCWP V-waves are determined by three major pathophysiological parameters: moderate or more severe MV regurgitation, LA dilatation, and elevated LVEDP.

## Introduction

With an aging population and improved treatment of acute and chronic cardiovascular diseases, the prevalence of congestive heart failure (CHF) and mitral regurgitation (MR) is increasing worldwide.^1^ MR and CHF are commonly causative of and significantly contribute to each other’s pathophysiology and manifestation.^2,3^ Shared disease process and similarities in presentation demand nuanced evaluation of MR in the setting of CHF to provide optimal medical, interventional, and surgical management.

When suspected on clinical grounds, modern day MR quantification relies on echocardiographic evaluation aided by invasive hemodynamic assessment when clinical and non-invasive findings conflict.^3,4^ Increased pulmonary capillary wedge pressure (PCWP) V-wave amplitude has been historically associated with severe MR,^3^ but while normal V-waves effectively rule out severe MR, increased V-waves do not necessarily rule it in.^5,6^ It has been suggested that elevated left ventricular end-diastolic pressure (LVEDP), a hallmark of CHF,^7^ may also contribute to increased V-wave amplitude.^8,9^ In addition, while significant chronic MR leads to left atrial (LA) dilatation,^10^ enlarged LA volume has been associated with increased V-waves in patients with^11^ and without significant MR.^12^

Despite of these controversies, there is paucity of modern studies exploring interaction between MR, elevated LVEDP, and increased LA volume in the genesis of increased PCWP V-waves. To fill this knowledge gap, this study was conducted to evaluate and compare the effects of MR, LVEDP, and LA volume on the PCWP V-wave amplitude in clinically stable, euvolemic CHF patients on guideline-directed therapy (GDT) undergoing elective left and right cardiac catheterization for dyspnea evaluation.

## Materials and Methods

This was a single tertiary care center study. The study cohort included consecutive patients referred for left and right cardiac catheterization for evaluation of shortness of breath by the treating physicians during the calendar years 2018-2022. Patients with acute coronary syndromes (ACS), decompensated heart failure, non-MV valvular disease of moderate or worse severity, MV stenosis of mild or worse severity, mechanical valve, pericardial effusion, constriction, on mechanical ventilation, on mechanical support with intra-aortic balloon pump, Impella®, or left ventricular assist device were excluded from the study.

The study was approved by the Institutional Review Board (IRB). The data was collected prospectively for the first 76 patients. Interim IRB review waived the informed consent requirement, as the collected data included only what was required for routine clinical care and no experimental measurements or treatments were performed. Data on the remaining patients was collected retrospectively. Clinical, echocardiographic, and invasive hemodynamic data was obtained through chart review, hemodynamic tracing analysis, and echocardiographic image processing.

### Hemodynamic data acquisition

Cardiac catheterization was performed according to the standard of care utilizing sterile technique, with standard catheters to assess pulmonary and right and left cardiac chamber hemodynamics. Procedures were performed with a patient in a supine position. Transducers were leveled and zeroed prior to procedure with mid-chest used as a reference point. Hemodynamic data was collected according to the standard protocols, waveforms and values were verified to ensure adequate data acquisition. Upward deflection of the PCWP tracing following QRS was interpreted as a V-wave with maximum value measured during late ventricular systole. PCWP and V-waves were assessed at end expiration. LVEDP, PCWP, and V-wave values were recorded in millimeters of mercury (mmHg).

### Echocardiographic data acquisition

Complete transthoracic echocardiograms were acquired in the ICAEL-accredited non-invasive imaging laboratory by certified sonographers and permanently stored off-line for the subsequent interpretation. The median time interval between invasive hemodynamic assessment and echocardiographic evaluation was 4+/-5.7 days, in 124/153 (81%) patients within 4 weeks interval. Echocardiograms were interpreted by the National Board of Echocardiography (NBE) certified cardiologists independently from the study, at the time when the test was performed for clinical indications. MR, LV ejection fraction and filling pressure, and left atrial volume evaluation was performed in accordance with American Society of Echocardiography Recommendations.^4,13,14^ Specifically, left atrial volume was determined by 2D-biplane method based on maximal end-systolic left atrial areas obtained in the carefully selected apical four chamber and apical two chamber views.^13^ LA was defined as dilated if LA volume exceeded 50 ml.^13^ Transmitral peak E-flow velocity was measured in the apical 4 chamber view using pulsed wave Doppler and e’ tissue Doppler velocity was obtained from the apical 4 chamber view by placing pulsed Doppler sampling volume in the basal septal and left ventricular lateral myocardial segments.^14^

## Results

Demographics and clinical characteristics are noted in Table 1. Moderate or worse MR was present in 20% (Table 1) but more than 50% of the patients manifested with V-waves greater than 20 mmHg and in 27% of patients the V-wave exceeded PCWP by 7 mmHg or more (Table 1). V-wave amplitude was significantly increased in patients with moderate or more significant MR (Figure 1, Panel A, p<0.001) or with dilated LA (Figure 1, Panel B, p=0.002). There was a trend towards increased V-wave amplitude in patients with elevated LVEDP (Figure 1, Panel C, p=0.117).

**Table 1.** Demographic, clinical, and hemodynamic characteristics.

| Characteristic | Value |
| --- | --- |
| Age, mean – years old | 68.4 +/- 11.9 |
| Male sex — no. (%) | 64/153 (41.8) |
| Body Mass Index, mean – kg/m<sup>2</sup> | 29.4 +/- 7.0 |
| Coexisting condition — no. (%) |  |
| Coronary artery disease | 97/153 (63.4) |
| Coronary artery bypass graft surgery | 29/153 (18.9) |
| Hypertension | 113/153 (73.9) |
| Diabetes | 54/153 (35.3) |
| Atrial fibrillation | 42/153 (27.5) |
| Ventricular pacemaker | 9/153 (5.9) |
| Hemoglobin, mean – g/dL | 12.6 +/- 2.2 |
| Brain natriuretic peptide, mean – pg/ml | 942 +/- 1103 |
| Glomerular filtration rate, mean – ml/min | 72.9 +/- 42.0 |
| Medical Therapy — no./total no. (%) |  |
| Beta-adrenergic antagonists | 92 (60.1) |
| ACE-I or ARB, or ARNI, or Hydralazine and Nitrates | 106 (69.3) |
| Aldosterone antagonists | 16 (10.4) |
| Loop diuretics | 68 (44.4) |
| Thiazides | 14 (9.2) |
| Systolic blood pressure, mean – mmHg | 123.4 +/- 25.8 |
| Diastolic blood pressure, mean – mmHg | 65.9 +/- 17.2 |
| Heart rate, mean – beats per minute | 80.2 +/- 16.4 |
| Left ventricular ejection fraction, mean – % | 46.2 +/- 15.9 |
| Left ventricular ejection fraction < 35 % — no. (%) | 45/153 (29.4) |
| Left atrial volume, mean – ml | 72.3 +/- 29.3 |
| Left atrial volume >50ml, no. (%) | 116/153 (75.8) |
| Left atrial volume indexed by BSA, mean – ml/m<sup>2</sup> | 36.1 +/- 14.6 |
| Left atrial volume indexed by BSA >34ml/m<sup>2</sup>, no. (%) | 72/153 (47.1) |
| Mitral regurgitation, moderate or more severe, no. (%) | 31/152 (20.4) |
| E/e', mean | 3.880 +/- 2.015 |
| Cardiac output, mean – L/min | 6.0 +/- 1.7 |
| Cardiac index, mean – L/min/m<sup>2</sup> | 3.0 +/- 0.7 |
| Left ventricular end-diastolic pressure (LVEDP), mean – mmHg | 13.5 +/- 8.7 |
| Left ventricular end-diastolic pressure >12 mmHg, no. (%) | 58/126 (46.0) |
| Mean pulmonary artery pressure, mean – mmHg | 19.8 +/- 8.3 |
| Pulmonary capillary wedge pressure (PCWP), mean – mmHg | 15.9 +/- 8.6 |
| V-wave, mean – mmHg | 22.1 +/- 11.7 |
| V-wave >20 mmHg, no. (%) | 81/150 (54) |
| V-wave and mean PCWP difference >7mmHg, no. (%) | 40/150 (26.7) |

**Figure 1.**
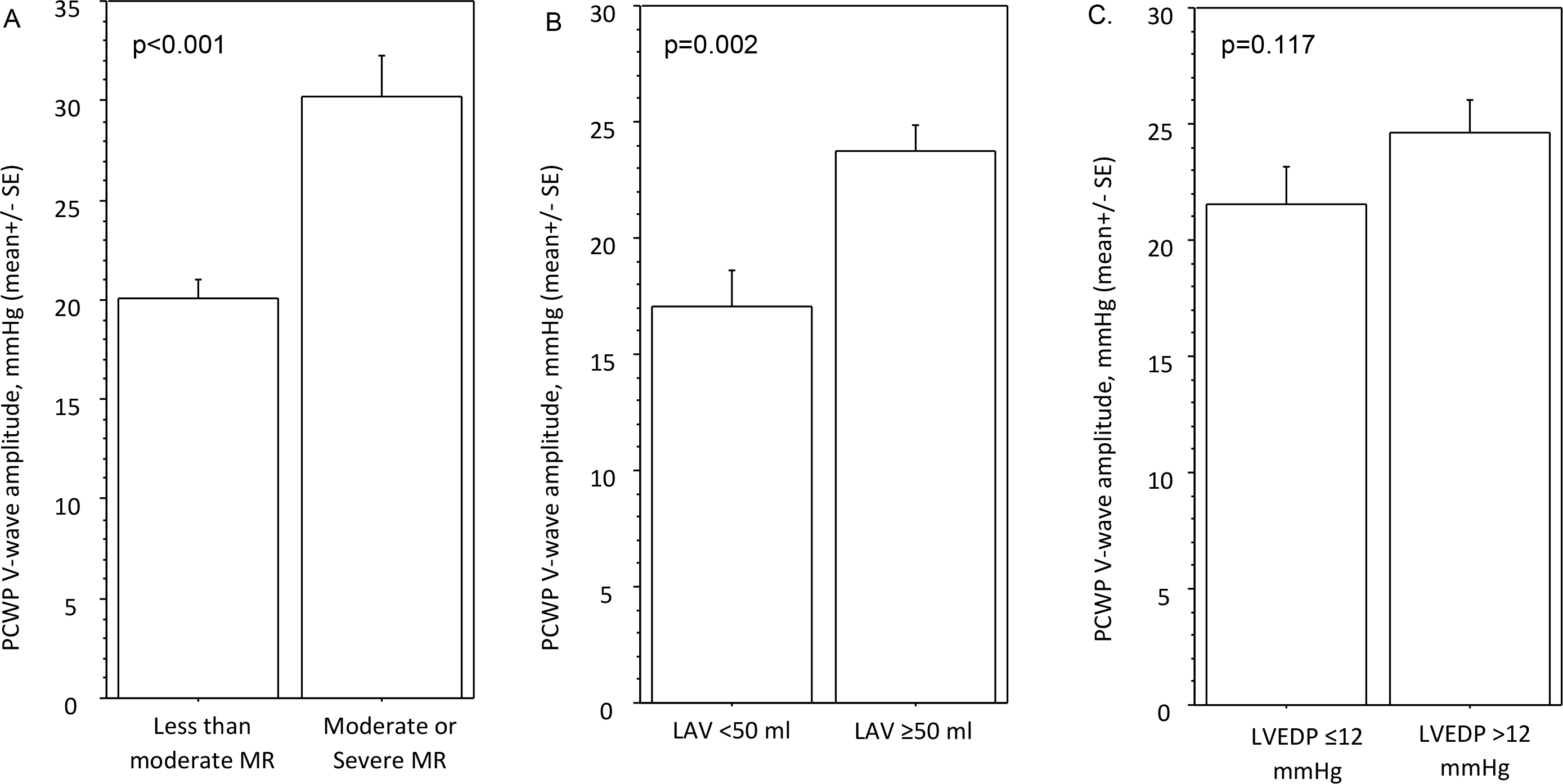
Effects of mitral regurgitation, left atrial volume, and LVEDP on PCWP V-wave amplitude. V-wave amplitude was significantly increased in patients with moderate or more significant MR (Panel A, p<0.001) or dilated LA (Panel B, p=0.002). There was a trend towards increased V-wave amplitude in patients with elevated LVEDP (Panel C, p=0.117).

Crosstabulation of the cohort according to the LA volume and degree of MR revealed three distinct patient populations, (1) patients with normal LA volume with less than moderate MR (n=36/153, 26%), (2) patients with dilated LA and less than moderate MR (85/153, 54%), and (3) patients with moderate to severe MR (31/153, 20%) with all but one with dilated LA. Within these three patient groups, when compared to patients with less than moderate MR and with normal LA volume, LVEDP was significantly elevated (Figure 2, panel A, p=0.039) and V-waves exceeding PCWP by 7 mmHg were significantly more common in patients with moderate or more severe MR and dilated LA (Figure 2, panel B, p=0.002).

**Figure 2.**
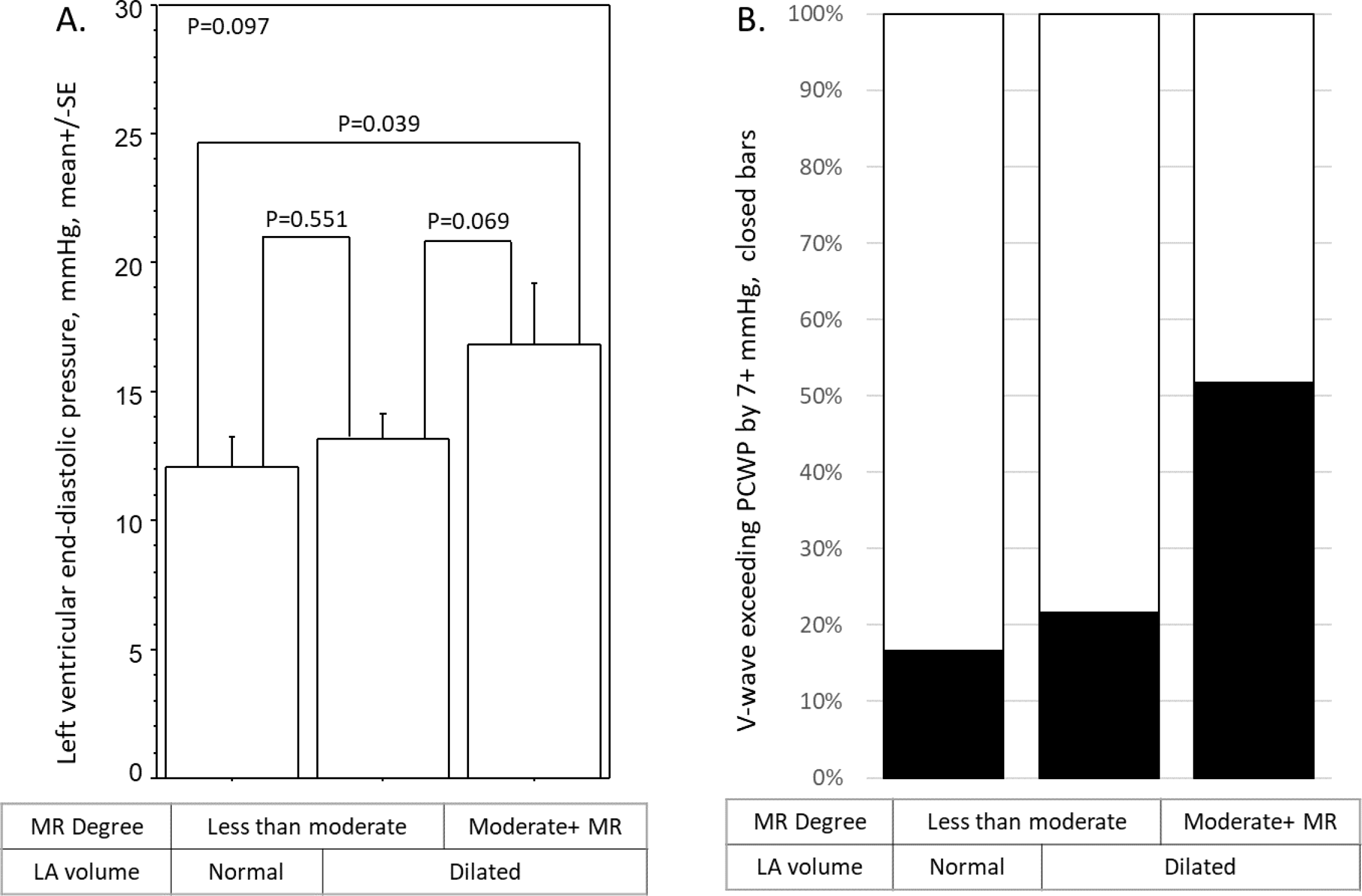
Effects of mitral regurgitation and left atrial size on LVEDP and prevalence of V-waves exceeding PCWP by 7 mmHg. Moderate or more severe MR and dilated LA were associated with elevated LVEDP (Panel A, p=0.039) and increased prevalence of V-wave exceeding PCWP by 7 mmHg (Panel B, p=0.002).

To analyze effects of LA volume, degree of MR and LVEDP on V-waves, the cohort was then subdivided into 6 anatomical, functional, and hemodynamic profiles according to the LA volume, degree of MR, and LVEDP (Table 2). Demographic and clinical characteristics were very similar between these six profiles, except for higher E/e’ ratio and lower hemoglobin level in patients with dilated LA, significant MR, or elevated LVEDP (Table 2, p<0.001 and p=0.008, respectively). Consistent with a compensated volume status required for study enrollment, average E/e’ ratio was significantly less than 14 in all patient profiles (Table 2).

**Table 2.**
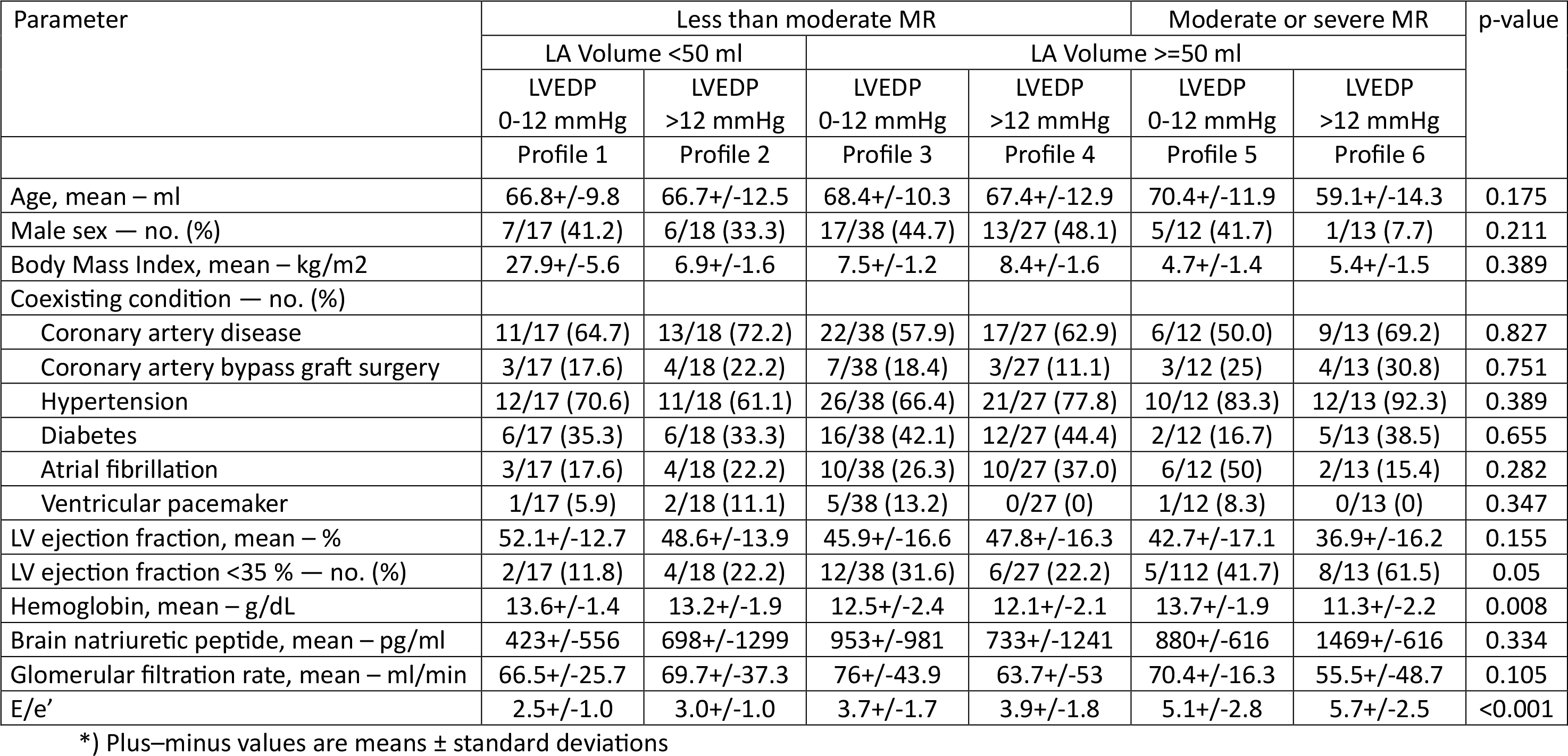
Demographic, clinical, and hemodynamic characteristics according to the structural and functional categories.

Effects of LA volume, degree of MR, and LVEDP on prevalence of elevated V-waves were highly significant (Figure 3, p<0.002). When the MR degree was moderate or worse, LA was dilated, and LVEDP was elevated, increased >20 mmHg V waves were present in all patients (Figure 3, profile 6). V waves >20 mmHg were also noted in two-thirds of patients with dilated LA and elevated LVEDP or if MR of moderate or worse in severity, but with a normal LVEDP (Figure 3, profiles 4 and 5). Finally, V waves > 20 mmHg were present in half of the patients with less than moderate MR and dilated LA or elevated LVEDP (Figure 3, profiles 2 and 3).

**Figure 3.**
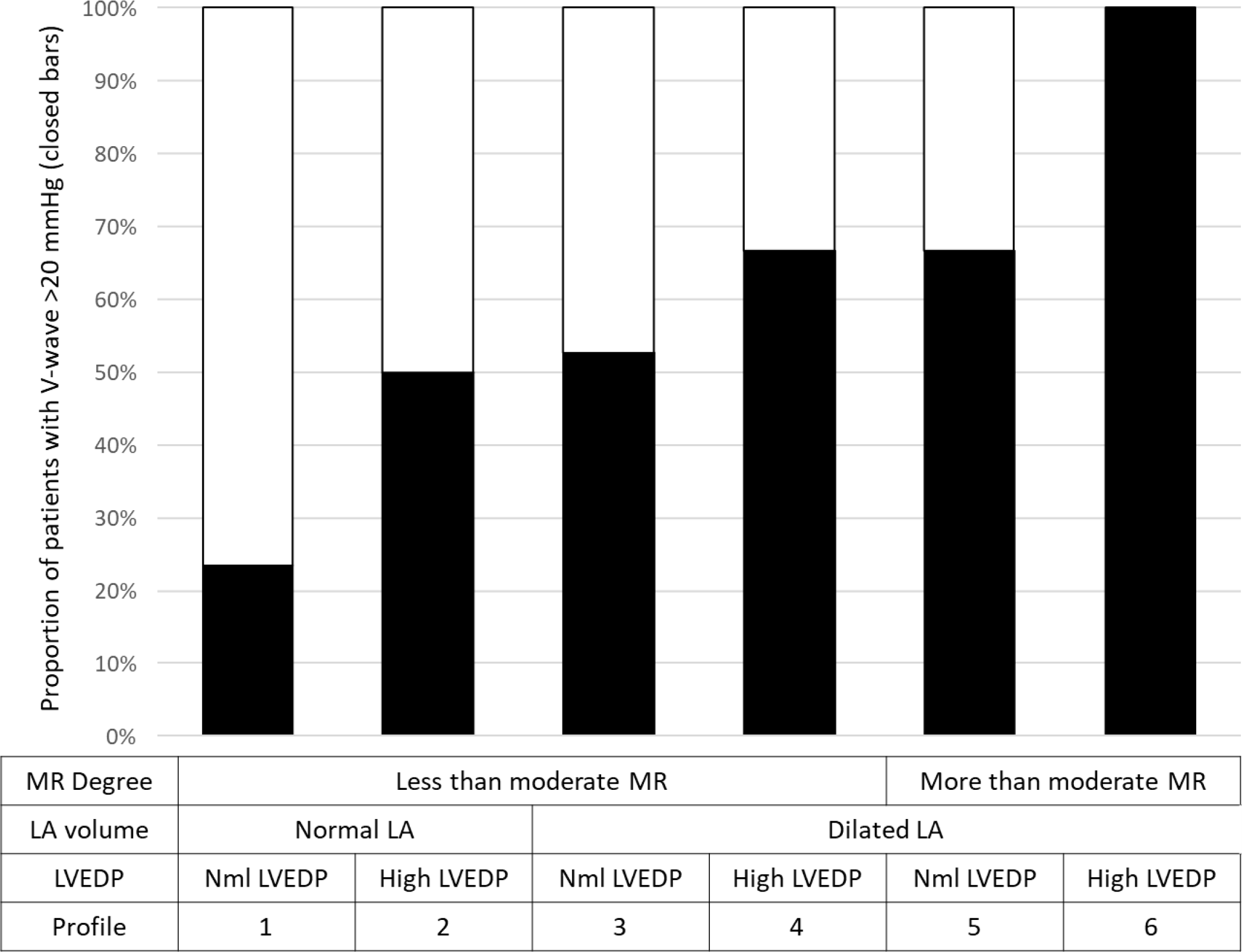
Effects of left atrial size, mitral regurgitation, and elevated LVEDP on PCWP V-wave amplitude LA volume, MR, and elevated LVEDP significantly affected prevalence of elevated V-waves (Panel A, p<0.002). In moderate or more severe MR with dilated LA and elevated LVEDP, V waves >20 mmHg were present in all patients (Panel A, profile 6). Elevated >20 mmHg V waves were also noted in two-thirds of patients with dilated LA and elevated LVEDP or if MR was moderate or more severe, but LVEDP was normal (Panel A, profiles 4 and 5), and in half of the patients with less than moderate MR but dilated LA or with elevated LVEDP (Panel A, profiles 2 and 3).

## Discussion

The prevalence of MR and CHF is increasing worldwide.^1^ Both MR and CHF manifest with shortness of breath and pulmonary congestion caused by synergistic disease processes leading to decreased LV and LA compliance with increased LV, LA, and pulmonary capillary wedge pressures.^2,3^ Optimal medical, interventional, and surgical outcomes in patients with co-existing MR and CHF demand a thorough understanding of cardiac hemodynamics in this complex disease process. While modern diagnosis and quantification of MR relies on non-invasive echocardiographic evaluation, left and right cardiac catheterization is employed when noninvasive testing results are inconclusive or conflict with clinical findings.^2,3^

PCWP V-waves reflect a rise in the LA pressure in late systole. Increased V-waves on PCWP tracing have been classically attributed to significant MR,^3^ however, clinical practice and prior research suggest that increased V waves may also occur without significant MR.^5,6,8,9^ Our study of stable euvolemic patients revealed three major contributors to increased PCWP V-wave amplitude, i.e. rise in the LA pressure in late systole: anatomical - LA dilatation, functional - MR of moderate or worse severity, and hemodynamic - elevated LVEDP synonymous with elevated diastolic LA pressure. Increased V-waves were unlikely when all these parameters are normal. Alternatively, we have observed increased V-waves in all patients when all three of these parameters were abnormal. Any combination of moderate or more severe MR, dilated LA, and elevated LVEDP was associated with 50-65% prevalence of increased V-waves.

Increased LV end-diastolic, LA, and pulmonary capillary wedge pressures with dilated LA are hallmarks of chronic CHF.^7^ Similarly, chronic hemodynamically significant MR imposes volume load on the LV and LA, with eventual LV and LA enlargement and rise in LVEDP, LA, and PCWP.^3^ Due to the similarities in the disease process, elevated LVEDP is not specific in diagnosis of CHF or MR, but rather reflects decreased LV and LA compliance.^2,3^ From the standpoint of central hemodynamic assessment, increased LVEDP and LA pressure are inferred when PCWP is elevated in the absence of significant mitral valve stenosis. Underscoring the importance of elevated LVEDP in genesis of increased V-waves, in our study of patients with less than moderate MR, increased V-waves were noted in 50% of patients with elevated LVEDP and normal LA volume and in 65% when elevated LVEDP was combined with dilated LA. It is important to note that our study included patients that were euvolemic and hemodynamically stable and treated with afterload reduction, beta-adrenergic antagonists, and diuretics. It is likely that volume overloaded CHF patients with elevated LVEDP would have an even higher prevalence of increased V-waves. From the clinical perspective, as normal LVEDP has been associated with reduced prevalence of increased V-waves even in patients with significant MR and/or dilated LA, improved hemodynamics and decreased V-waves should be expected with meticulous volume management and aggressive LVEDP lowering.

Dilated LA is universally seen in patients with advanced CHF and in chronic hemodynamically significant MR. LA dilatation reflects chronic volume and pressure load on the LA, which may be due to increased LVEDP in CHF, increased preload due to MR, or a combination of both.^2,3^ Understanding importance of abnormal LA structure and function in central hemodynamics changes attributable to CHF and MR continues to evolve.^10-12^ Historically, chronic LA dilatation was thought to decreased LA pressure rise due significant MR.^5^ However, recent studies suggested that chronic atrial dilatation and decreased LA compliance augment transmission of MR-related systolic pressure increase into pulmonary capillary bed causing increased V-wave amplitude.^10,12^ In our study, LA dilatation was associated with increased V-wave amplitude, especially in patients with co-existing MR of moderate or worse severity. Comparing effects of increased LVEDP and dilated LA on V-waves, prevalence of increased V-waves was approximately 50% if either LVEDP was elevated or LA was dilated, and 65% if LVEDP was elevated in the setting of dilated LA. This observation indicates that compliance in dilated LA is decreased and dilated LA is unable to accommodate any additional volume without significant rise in pressure, thus generating increased V-waves.

Interpreting V-waves may be difficult in patients with elevated PCWP, when the absolute V-wave amplitude may be less important than the difference between V-waves and mean PCWP.^5,9^ In patients with elevated PCWP subtracting mean PCWP from the V-wave amplitude promises to reduce effects of elevated LVEDP on V-waves. The absolute difference between peak V-wave and mean PCWP consistent with hemodynamically significant MR has not been established with the values above 5-7 mmHg generally considered to be meaningful.^5,9^ When V-waves exceeding LVEDP by 7 mmHg or more were considered, only 50% of our study patients with moderate or more severe MR had significantly increased V-waves. These findings underscore the limited sensitivity of V-waves in MR diagnosis and the important contribution of elevated LVEDP to the pathophysiology of increased V-waves.

### Study Limitations and Strengths

Our study findings are limited to euvolemic patients referred for cardiac catheterization when invasive assessment was requested for clinical indications. Prevalence of increased V-waves may be different in patients who are hemodynamically unstable, require mechanical circulatory support and have significant valvular disease other than mitral regurgitation. Furthermore, additional hemodynamic and pathologic parameters that may affect V-wave amplitude. These parameters and conditions may include atrial arrhythmias, ventricular conduction abnormalities and atrio-ventricular blocks, acute LV dysfunction in the setting of myocardial ischemia and non-ischemic conditions like myocarditis and stress cardiomyopathy.

We are unable to conclusively evaluate the effect of heart failure GDMT on V-waves. In our study, two-thirds of patients were on afterload reduction and beta-adrenergic antagonists, but the treatment was not randomized. In addition, many patients were on diuretics with a small fraction on aldosterone-receptor antagonists, and only a few on SGLT-2 inhibitors, as the data was collected before the wide-spread acceptance of this treatment modality. Likewise, we cannot speculate on effects of inotropes and/or mechanical left ventricular support on V-waves, which will require a separate study specifically aimed at investigating these extremely vulnerable patients.

With limitations as noted above, our findings are biologically plausible and reveal highly significant differences in the study endpoints despite a modest sample size. These findings contribute to and extend the body of knowledge on complex hemodynamic changes attributable to heart failure and mitral regurgitation.

## Conclusions

Our data indicates that in euvolemic patients undergoing elective invasive hemodynamic evaluation, increased V-wave amplitude is determined by three major pathophysiological parameters: 1) anatomy of LA, normal vs. dilated, 2) degree of MR, less than moderate vs. moderate or more severe, and 3) hemodynamics of LV, normal or elevated LVEDP. In our study, increased V-waves were very unlikely when all these parameters are normal. Any combination of 2 of these abnormal parameters was associated with 50-65% prevalence of increased V-waves and reached 100% when all three were abnormal. From the clinical perspective, these findings suggest that comprehensive interpretation of PCWP hemodynamic data should include anatomy of the left atrium, mitral valve function, and LV filling parameters.

## Data Availability

NA

## References

1. Tsao CW, Aday AW, Almarzooq ZI, Anderson CAM, Arora P, Avery CL, Baker-Smith CM, Beaton AZ, Boehme AK, Buxton AE, et al. Heart Disease and Stroke Statistics-2023 Update: A Report From the American Heart Association. Circulation. 2023;147:e93-e621. doi:10.1161/CIR.0000000000001123

2. Heidenreich PA, Bozkurt B, Aguilar D, Allen LA, Byun JJ, Colvin MM, Deswal A, Drazner MH, Dunlay SM, Evers LR, et al. 2022 AHA/ACC/HFSA Guideline for the Management of Heart Failure: A Report of the American College of Cardiology/American Heart Association Joint Committee on Clinical Practice Guidelines. Circulation. 2022;145:e895–e1032. doi: 10.1161/CIR.0000000000001063

3. Otto CM, Nishimura RA, Bonow RO, Carabello BA, Erwin JP, 3rd, Gentile F, Jneid H, Krieger EV, Mack M, McLeod C, et al. 2020 ACC/AHA Guideline for the Management of Patients With Valvular Heart Disease: Executive Summary: A Report of the American College of Cardiology/American Heart Association Joint Committee on Clinical Practice Guidelines. Circulation. 2021;143:e35–e71. doi: 10.1161/CIR.0000000000000932

4. Zoghbi WA, Adams D, Bonow RO, Enriquez-Sarano M, Foster E, Grayburn PA, Hahn RT, Han Y, Hung J, Lang RM, et al. Recommendations for Noninvasive Evaluation of Native Valvular Regurgitation: A Report from the American Society of Echocardiography Developed in Collaboration with the Society for Cardiovascular Magnetic Resonance. J Am Soc Echocardiogr. 2017;30:303–371. doi: 10.1016/j.echo.2017.01.007

5. Fuchs RM, Heuser RR, Yin FC, Brinker JA. Limitations of pulmonary wedge V waves in diagnosing mitral regurgitation. Am J Cardiol. 1982;49:849–854. doi: 10.1016/0002-9149(82)91968-3

6. Snyder RW, 2nd, Glamann DB, Lange RA, Willard JE, Landau C, Negus BH, Hillis LD. Predictive value of prominent pulmonary arterial wedge V waves in assessing the presence and severity of mitral regurgitation. Am J Cardiol. 1994;73:568–570. doi: 10.1016/0002-9149(94)90335-2

7. Iskandrian AS, Segal BL, Hakki AH. Left ventricular end-diastolic pressure in evaluating left ventricular function. Clin Cardiol. 1981;4:28–33. doi: 10.1002/clc.4960040107

8. Pichard AD, Diaz R, Marchant E, Casanegra P. Large V waves in the pulmonary capillary wedge pressure tracing without mitral regurgitation: the influence of the pressure/volume relationship on the V wave size. Clin Cardiol. 1983;6:534–541. doi: 10.1002/clc.4960061104

9. Kageji Y, Oki T, Iuchi A, Tabata T, Ito S. Relationship between pulmonary capillary wedge V wave and transmitral and pulmonary venous flow velocity patterns in various heart diseases. J Card Fail. 1996;2:215–222. doi: 10.1016/s1071-9164(96)80044-3

10. Pape LA, Price JM, Alpert JS, Ockene IS, Weiner BH. Relation of left atrial size to pulmonary capillary wedge pressure in severe mitral regurgitation. Cardiology. 1991;78:297–303. doi: 10.1159/000174808

11. Pizzarello RA, Turnier J, Padmanabhan VT, Goldman MA, Tortolani AJ. Left atrial size, pressure, and V wave height in patients with isolated, severe, pure mitral regurgitation. Cathet Cardiovasc Diagn. 1984;10:445–454. doi: 10.1002/ccd.1810100505

12. Purga SL, Karas MG, Horn EM, Torosoff MT. Contribution of the left atrial remodeling to the elevated pulmonary capillary wedge pressure in patients with WHO Group II pulmonary hypertension. J Echocardiogr. 2019;17:187–196. doi: 10.1007/s12574-018-0410-8

13. Lang RM, Badano LP, Mor-Avi V, Afilalo J, Armstrong A, Ernande L, Flachskampf FA, Foster E, Goldstein SA, Kuznetsova T, et al. Recommendations for cardiac chamber quantification by echocardiography in adults: an update from the American Society of Echocardiography and the European Association of Cardiovascular Imaging. J Am Soc Echocardiogr. 2015;28:1–39 e14. doi: 10.1016/j.echo.2014.10.003

14. Nagueh SF, Smiseth OA, Appleton CP, Byrd BF, 3rd, Dokainish H, Edvardsen T, Flachskampf FA, Gillebert TC, Klein AL, Lancellotti P, et al. Recommendations for the Evaluation of Left Ventricular Diastolic Function by Echocardiography: An Update from the American Society of Echocardiography and the European Association of Cardiovascular Imaging. J Am Soc Echocardiogr. 2016;29:277–314. doi: 10.1016/j.echo.2016.01.011

